# The Citation of Retracted COVID-19 Papers is Common and Rarely Critical

**DOI:** 10.1101/2022.06.30.22277084

**Authors:** Gideon Meyerowitz-Katz, Praba Sekhar, Lonni Besançon, Tari Turner, Steve McDonald

## Abstract

**Background:** Retraction is the final safeguard against research error/misconduct. In principle, retraction exists to prevent serious issues identified in published research through post-publication review. Our study investigated the citing of clinical research papers retracted during the COVID-19 pandemic.

**Methods:** We used the Retraction Watch database extracted as of 27/01/2022 to identify retracted COVID-19 papers and the Google Scholar citation function to gather a dataset of citations of retracted clinical research. We reviewed key aspects of the citing research.

**Results:** In total, the Retraction Watch database included 212 entries for retracted COVID-19 papers. Of these, 53 papers were clinical. There were a total of 1,141 citations of retracted papers, with 105 errors, leaving 1,036 citations to analyze. The majority (86%) of citations were not critical. The majority (80%) of papers citing retracted research were published after the retraction date.

**Conclusions:** The citation of retracted and withdrawn COVID-19 clinical studies is common, and rarely critical. Most researchers who cite retracted research do not identify that the paper is retracted, even when submitting long after the paper has been withdrawn. This has serious implications for the reliability of published research and the academic literature, which need to be addressed.

## Introduction

Retraction is the final safeguard against academic error and misconduct, and thus a cornerstone of the entire process of knowledge generation. In principle, retraction exists to prevent serious issues and inadequacies identified in published research through post-publication review from continuing to pollute the literature and having an ongoing erroneous impact on the development of scientific ideas (1). Papers can be retracted for a variety of reasons, including errors made by the authors that were only noticed after publication, mistakes in peer-review, and more serious cases of malfeasance such as plagiarism, fraud, and data fabrication, but ultimately the message is the same: this research is not trustworthy, and should not be used as a source of knowledge. There are very few mechanisms in academic literature through which studies can be reconsidered post-publication, making retraction a vital aspect of the scientific landscape.

Even prior to the COVID-19 pandemic, there was evidence that the retraction process was error-prone and fraught with misunderstanding. For example, papers that have been withdrawn from publication due to concerns about the “analysis of the data reported” (2) may still be cited years later without any discussion of the retraction (3). The retraction process itself is notoriously clumsy, inept, and slow-paced, with journals often taking years to act on even the most obvious and blatant issues in the research they publish (4). When a junior researcher noted obvious and extensive plagiarism of his work in the editorial of a more senior editor, the journal in question took more than a decade to act. The paper was finally withdrawn for “unlawful and indefensible breach of copyright” (5). The impact of retracted papers on scientific knowledge and clinical practice can continue well after they are retracted. A preprint review has shown that systematic reviews and meta-analyses which include research which is later retracted are rarely, if ever, updated once the issues with the included research come to light, while nearly half of the reviews citing retracted papers were published after those papers were retracted (6). Key recommendations for proper retraction processes remain largely unimplemented across the scientific literature (7).

Many issues with the research production and publication process were heightened during the COVID-19 pandemic, which has triggered an enormous explosion in academic literature. The surge in the publication of papers about COVID-19 resulted in more than 200,000 papers indexed by PubMed in the 20 months after the virus was first identified (8, 9). The expedited production and promulgation of COVID-19 papers has been well-documented, and has predictably resulted in a large number of errors throughout the process, which in turn have resulted in retractions (10). As of June 2022, the website Retraction Watch has chronicled 233 retracted papers relating to COVID-19, with the number growing steadily.

Prior research has identified serious deficiencies in the retraction process during the pandemic. A cross-sectional review published in 2021 found that the reason for retraction was commonly omitted from retraction notices, and that more than half of retracted COVID-19 papers remained available as an original document without indication that the paper had been removed (11). A notable study of cardiovascular disease and COVID-19 which was retracted in June 2020 one month after it was published, was cited more than 600 times (12) in the following year and was still being repeatedly, uncritically cited 11 months later.

This leads to an important question – when retractions occur, are they noticed by the scientific community? If retraction is a guard against the incorporation of low-quality, error-filled, wrong or even fake research, is it having an impact on the use of papers as evidence in future scientific research? If not, it suggests that the final bastion of rigor in academic publishing is not only flawed, but potentially useless as a mechanism for removing poorly-conducted, mistaken, or even fraudulent research from the literature.

We conducted a review of clinical COVID-19 papers retracted during the pandemic to investigate the citation of those papers, and whether retraction prevents scientists using withdrawn studies.

## Methods

We used the Retraction Watch database extracted as of 27/01/2022 to identify retracted COVID-19 papers. This database is maintained by the Retraction Watch website, and is to our knowledge the most complete database of papers that have been withdrawn by either preprint servers or journals during the pandemic, and includes the name, date of publication, date of retraction, and article type of the papers in question.

To identify how many citations each retracted paper had received, we used the Google Scholar ‘citation’ function. This is a weblink present below each paper when searched using Google Scholar. There are known issues with the Google Scholar citation count, as it has high sensitivity but low specificity, however this was considered to provide the best coverage of potential retractions. We extracted the number of citations per retracted paper (RP) (13).

Using Google Scholar, we extracted data on papers that cited the RPs, called citing papers (CP). For CPs, we identified the date of publication, date of submission (where available), whether the CP cited the RP critically (i.e. identifying that the paper was retracted, giving reasons for retraction), whether the CP was a systematic review including the RP, the publishing journal, and the URL. The dates of publication and retraction were used to calculate the number of days in between the publication/submission of the CP and the date of retraction of the RP for each case.

After determining the total number of citations for each RP, we excluded non-clinical research (i.e. editorials, perspectives, etc) from further data extraction. There were two reasons for this: firstly, non-clinical research was considered to be less important to the pandemic response than clinical studies, systematic reviews, and similar. Secondly, the median and average number of citations that non-clinical retracted research received was far lower, with a median of 3 vs 12 citations and an average of 18 vs 80 per paper. Therefore, we summarized this research descriptively, but did not extract citation information on the CPs for non-clinical RPs.

We then further divided the papers into those which had more or fewer than 100 citations total. For papers with more than 100 citations (n=9), we reviewed only the first 100 citations given by Google Scholar’s “sorted by relevance” category. Since papers with the most citations appear first, we used the first 100 citations as an approximation of the most impactful papers citing these RPs.

We excluded from our analysis papers that were only temporarily retracted.

All statistical analysis was conducted in Stata 15 and Excel. Where papers were published in languages other than English, Google Translate and DeepL were used to determine whether the citation was critical.

## Results

In total as of 27/01/2022, the Retraction Watch database included 212 entries for retracted papers. The categories are summarized below in Table 1:

**TABLE 1.**
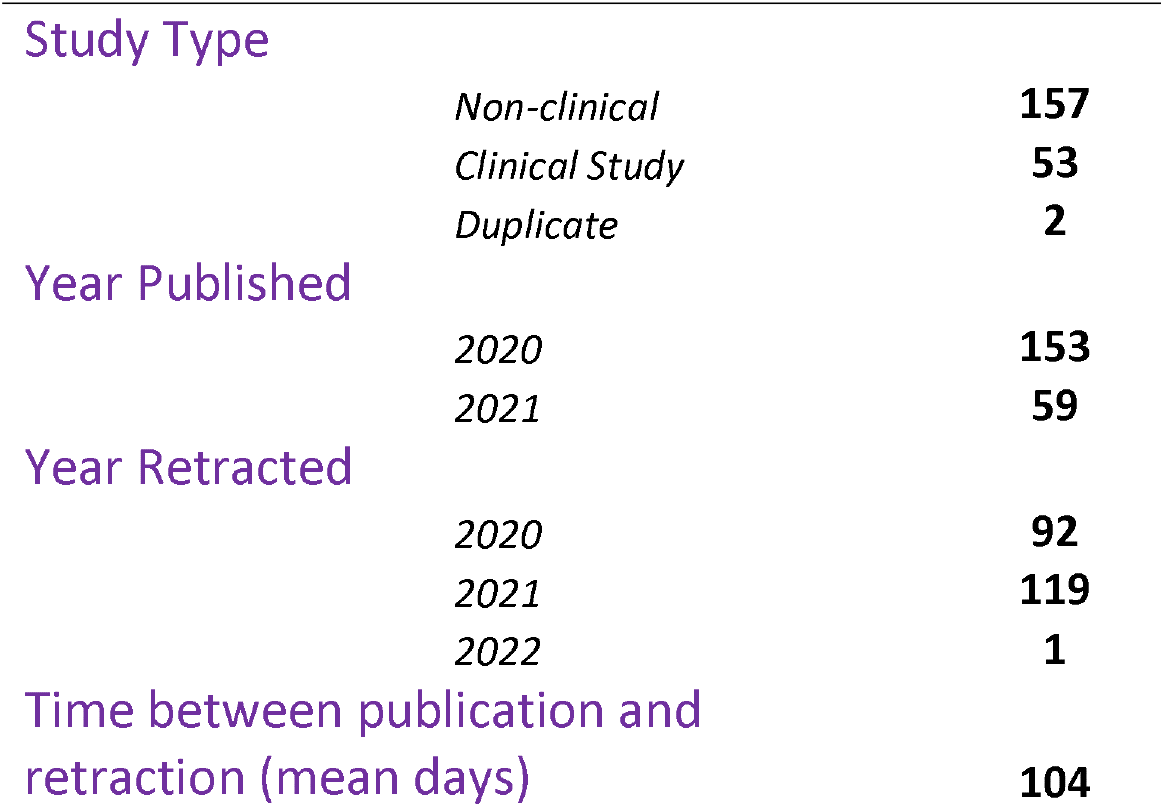

Study Type
|  |  |
| --- | --- |
| *Non-clinical* | **157** |
| *Clinical Study* | **53** |
| *Duplicate* | **2** |
| Year Published |  |
| *2020* | **153** |
| *2021* | **59** |
| Year Retracted |  |
| *2020* | **92** |
| *2021* | **119** |
| *2022* | **1** |

The dates of RPs varied. 72% of RPs were published in 2020, while 28% were published in 2021. There were 2 duplicates identified in the dataset, as well as 4 papers that were retracted but ultimately republished. There were 53 clinical studies that were retracted, not duplicates, and never republished which met criteira for inclusion in our analysis.

Analyzed papers had a median of 7 citations, and a mean of 53. The modal number of citations was 0, with a total of 2,697 citations. The most cited paper with 1,360 citations at the time of data extraction was *“Hydroxychloroquine or chloroquine with or without a macrolide for treatment of COVID-19: a multinational registry analysis” (14)* published in May 2020 in The Lancet, and retracted two weeks later in early June.

We extracted a total of 1,141 citations of RPs. Of these citations, 105 were either inaccessible papers, duplicate citations, or errors introduced by Google Scholar (i.e. this paper (15) which did not cite any COVID-19 research at all) leaving 1,036 citations to review.

The vast majority of CPs were not critical, with 893/1,036 (86%) not identifying that the RP had been retracted or raising any concerns about the research in the text. The remaining 143/1,036 (14%) citations were critical, either explicitly mentioning that the research was retracted or noting that there were reasons for questioning the reliability of the RP. An example of a critical citation is an update to a systematic review that excluded a paper after it had been retracted (16).

However, the proportion of critical citations was exaggerated by a small number of infamous studies. The most cited paper (14) was responsible for 62/143 (43%) of the total critical citations. When this paper was removed from the analysis, the proportion of critical citations dropped even further, with 91% of all CPs not mentioning the fact that the RP was retracted or noting any concerns with the quality of that research.

There were 812 papers with sufficient information to extract date of publication as well as date of retraction of the RP. The majority (80%) of these CPs were published substantially after the RP was retracted, with a median of 133 days post retraction (Figure 1). One paper was published a full 635 days after the RP it cited was retracted. Critical CPs were published a similar length of time after retraction to non-critical CPs, with a median of 110.5 (critical) days compared to 138 (non-critical), p=0.159 when compared using a Kruskal-Wallis test.

**Fig 1.**
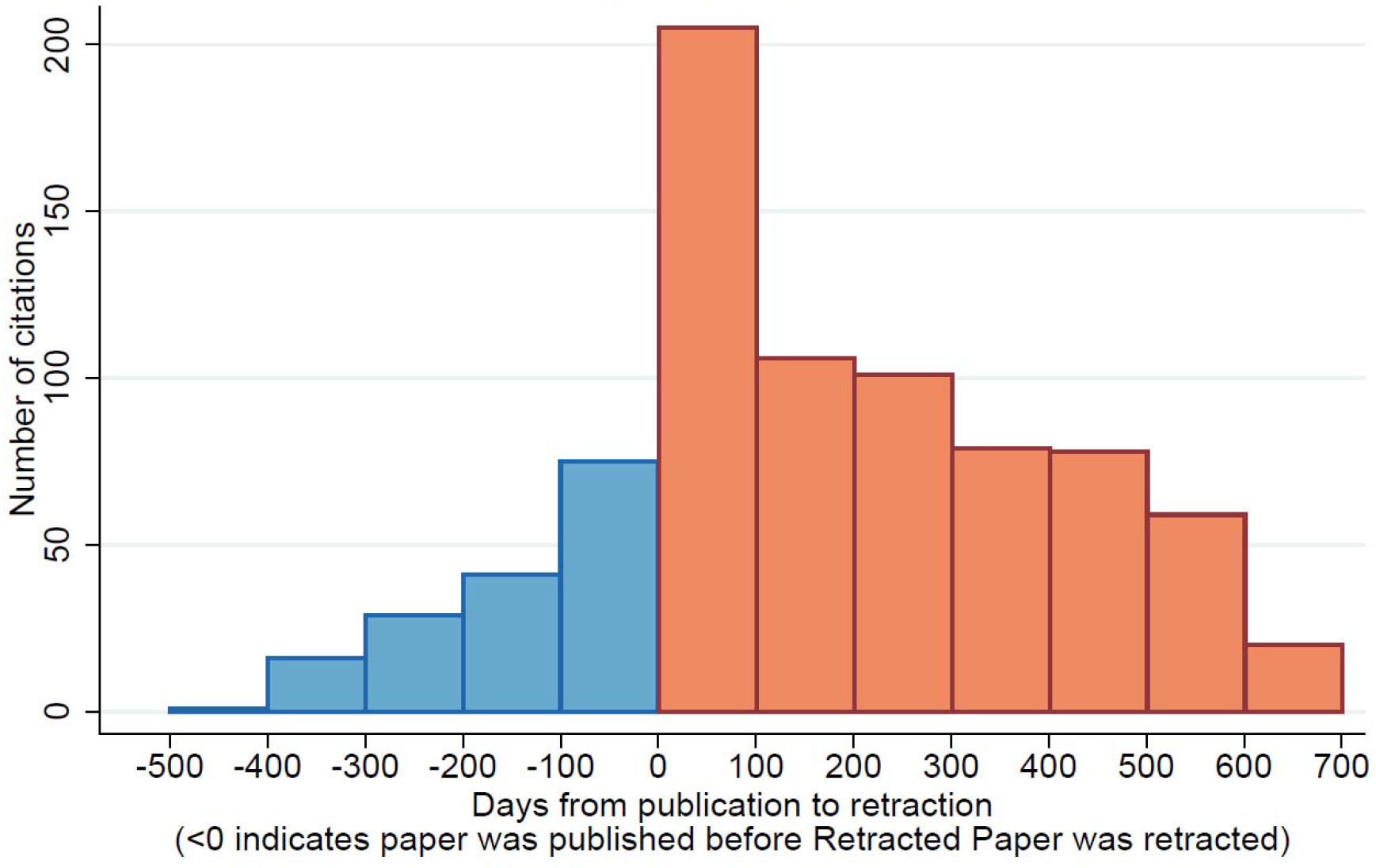
Histogram of times from Retracted Paper retraction to Citing Paper publication

A subset of citing papers also had information on the submission date of the paper available online. Of these 489 papers, the slight majority (52%) were submitted before the RP was retracted, with a substantial minority (48%) submitted after the RP had already been withdrawn (Figure 2). In the most extreme case, a paper (17) which cited a piece of retracted research (18) was submitted a full 640 days after the paper had been withdrawn, and was uncritical in this citation.

**Fig 2.**
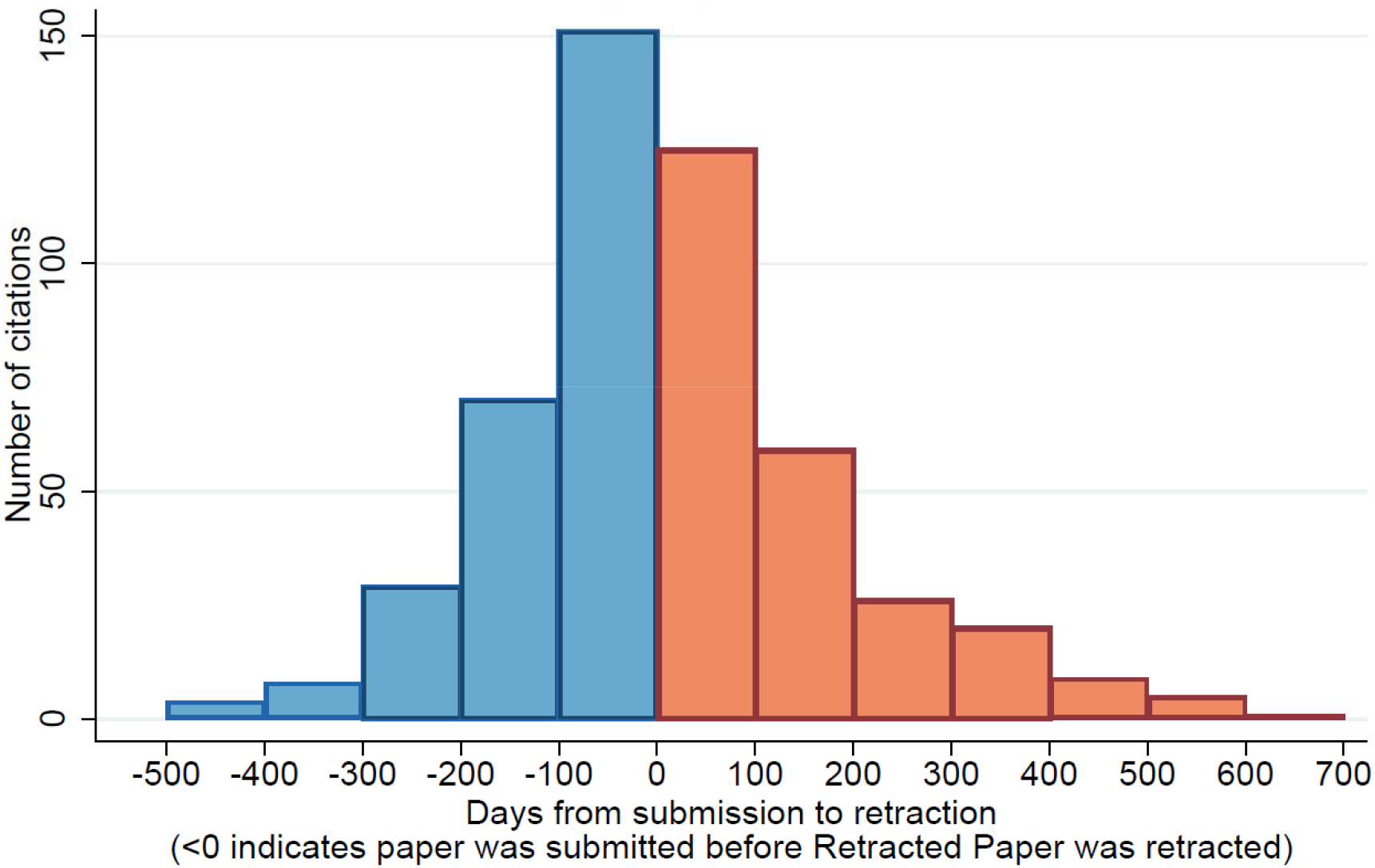
Histogram of times from Retracted Paper retraction to Citing Paper submission

About a quarter (26%) of critical citations were in commentaries, which often were written specifically due to the RP and focused on this paper exclusively. 12% of critical citations were meta research papers explicitly aimed at reviewing retracted research. A further 10% were some sort of editorial announcement pertaining to the RP, such as an expression of concern or a notice of retraction. Only 43% (62/143) of critical citations were either original research or review papers that clearly noted the RP was poor-quality or withdrawn.

## Discussion

This study highlights a critical flaw in research production, publication, and review during the COVID-19 pandemic. The citation of retracted and withdrawn COVID-19 clinical studies is common, and very rarely critical. Most researchers who cite retracted research do not note that the paper is retracted, even when submitting long after the paper has been withdrawn. When publications do note that research has been retracted it is often in the context of meta-research of research integrity or editorial announcement noting the retraction. This calls into question the value of retraction as a mechanism for removing problematic research from the literature, and shows a serious flaw in a large portion of COVID-19 research that may have directly impacted patient care.

This issue is even more concerning because many of the already rare, critical citations of retracted research appear to be due to high-profile papers becoming infamous for their retraction. The Surgisphere scandal, involving The Lancet and the New England Journal of Medicine, was publicized across the world. The retracted paper with both the most individual citations and the most critical citations (14) was part of this scandal, and was clearly notorious *because* it was retracted. This paper made up nearly half of all critical citations, and was one of the few papers cited more often critically than uncritically. In this dataset not only was critical citation of retracted research rare, but that critical citation was often predicated on the infamy of the retraction itself.

Some of the retractions, and the subsequent lack of identification of these as being retracted in citing studies, may result from expedited publication processes aiming to increase access to evidence to guide practice during a pandemic. Publication processes certainly have been accelerated for COVID-19 research; time to acceptance of publications was more than 11 times faster for COVID-19 publications than influenza publications in 2020 (19). While rapid translation of research into practice is vital in the context of a new and widespread disease; this research highlights that efficiency gains which come at the cost of reduced quality control may have long-term negative impacts on research and practice.

However, it is also notable that these retracted papers were – by definition – extremely low-quality even before being withdrawn. While the citation of unreliable research may in part be blamed on the rush to publish, there also appears to be a notable lack of rigor in the citations generally. For example, one retracted paper (20) was cited many times prior to the removal of the manuscript online despite impossible values and extremely poor methodology in the report itself (21). Even if authors cannot be expected to notice clear signs of fabrication in the research they cite, it is perhaps a problematic sign that even overt indications of low-quality science fail to prevent papers from being cited.

The burgeoning role of preprints in dissemination of research has implications for both increasing speed of access to research results, and increasing opportunity for promulgation of unreliable research through side-stepping of peer review. Preprints have made up nearly one third of COVID-19 research publications (22, 23) and have been a vital part of ensuring rapid access to the evidence about effective treatment of COVID-19. However accepted standards and processes for use of preprint data for informing clinical and research decisions are still in development and methods for retraction of preprints are just one area that needs further exploration.

It would be unfair though to single out preprints as the main source of retracted research. As noted above, the most high-profile retracted papers were originally published in some of the world’s most well-respected journals. Preprints may be one avenue for unscrupulous researchers to pursue publication of their low-quality or fraudulent research, but it is clear that academic journals have been just as culpable in the promulgation of problematic papers despite the hurdle of peer review.

One issue that has been noted previously is the inadequate and incomplete nature of many retractions. Of retracted papers, many are left without overt signs that the manuscript has been withdrawn (11), while still further manuscripts are retracted but with little information given as to the reason for retraction even if the paper has been determined to be entirely fraudulent (24). This may in part explain the ubiquitous citation of retracted research – authors who are acting in good faith are not aware that the papers have been retracted, or believe that the reason for retraction does not undercut the reason that they have cited the research. This is a well-understood problem, with already-proposed solutions to reduce the inadvertent citation of retracted research (7).

Unfortunately, this cannot explain the entire problem. There is clearly a large portion of the research community that does not assess the reliability or retraction status of the papers that they cite. One potential avenue to contribute to addressing this would be an automated system within publication software allowing for an automatic flag to the editorial team of citation of retracted research.

There are two main limitations to the strategy that we pursued for this paper. Firstly, we only reviewed the first 100 citations of retracted papers with >100 total citations. While this made up a small minority of the total RPs, it did consist of a large proportion of the total citations of retracted research. It is plausible that analyzing all citations would change our results, although given the consistency with which authors cite RPs we would not expect this to have a substantial impact.

Secondly, we used the ‘most relevant’ function on google. The algorithm underlying this is unclear but appears to reflect the highest-quality citing papers (13), and specifically seems to weight the number of citations that citing papers receive heavily. In other words, those papers which are more commonly cited appear higher in the ‘most relevant’ category. For most papers, this is likely to have had no impact on the results, but it may have changed the results for papers with >100 total citations.

One final limitation of our paper was that we only considered clinical research, leaving aside perspectives, commentaries, letters to the editor, editorials, and other retracted non-clinical research. We chose to do this as we felt that clinical papers had the most relevance to patient care during the pandemic, and also represented the vast majority of all citations of retracted research. It is however possible that non-clinical research does not follow the patterns we have uncovered in this paper.

## Conclusions

The citation of retracted and withdrawn COVID-19 clinical studies is common, and very rarely critical. Most researchers who cite retracted research do not identify that the paper is retracted, even when submitting their paper long after the retracted paper has been withdrawn. This has serious implications for the reliability of published research and the academic literature as a whole. Action is needed to minimise the impact of retracted research studies on future research and clinical practice in COVID-19 and beyond.

## Data Availability

Data on retractions is available on request from Retraction Watch. Citations data is available on request from the corresponding author once retraction data has been shared by RW.

## DECLARATION OF INTEREST

The authors declare no conflicts of interest for this research.

## CRediT author statement

**Gideon Meyerowitz-Katz:** Conceptualization, Methodology, Formal analysis, Data Curation, Writing – Original Draft, Writing – Review & Editing. **Praba Sekhar:** Data Curation, Writing – Review & Editing. **Lonni Besançon:** Data Curation, Writing – Review & Editing. Tari Turner: Methodology, Data Curation, Writing – Review & Editing. **Steve McDonald**: Methodology, Data Curation, Writing – Review & Editing.

## FUNDING

This research was not funded.

## Notes

### Competing Interest Statement

The authors have declared no competing interest.

